# Antibodies in Serum and MENSA Predict Non-recurrence in Primary *Clostridioides difficile* infection

**DOI:** 10.1101/2021.11.04.21265901

**Authors:** Natalie S. Haddad, Sophia Nozick, Geena Kim, Shant Ohanian, Colleen S. Kraft, Paulina A. Rebolledo, Yun Wang, Hao Wu, Adam Bressler, Sang Nguyet Thi Le, Merin Kuruvilla, L. Edward Cannon, F. Eun-Hyung Lee, John L. Daiss

## Abstract

**Background:** Within eight weeks of primary *Clostridioides difficile* infection (CDI), as many as 30% of patients develop recurrent disease with the associated risks of multiple relapses, morbidity, and economic burden. There are no validated biomarkers predictive of recurrence during primary infection. This study demonstrates the potential of a simple test for identifying hospitalized CDI patients at low risk for disease recurrence.

**Methods:** Forty-six hospitalized CDI patients were enrolled at Emory University Hospitals. Serum and MENSA samples prepared during weeks 1, 2, and 4 following symptom-onset were measured for antibodies specific for ten *C. difficile* antigens.

**Results:** Among the 46 *C. difficile*-infected patients, nine (19.5%) experienced recurrence within eight weeks of primary infection. Among the 37 non-recurrent patients, 23 had anti-*C. difficile* MENSA antibodies specific for any of the three toxin antigens: TcdB-CROP, TcdBvir-CROP, and/or CDTb. Positive MENSA responses occurred within the first week post-symptom onset, including six patients who never seroconverted. A similar trend was observed in serum responses, but they peaked later and identified fewer patients (19/37). In contrast, none of the patients who subsequently recurred after hospitalization produced antibodies specific for the three *C. difficile* toxin antigens. IgA antibodies for the toxin antigens demonstrated the greatest predictive power for protection from recurrence.

**Discussion:** The development of IgG and/or IgA antibodies for three *C. difficile* toxins in serum and/or MENSA has prognostic potential. These immunoassays measure nascent immune responses that reduce the likelihood of recurrence. Early identification of patients at-risk for recurrence can reduce costs and morbidity.

## INTRODUCTION

*Clostridioides difficile* infection (CDI) is the most common cause of hospital-acquired infectious diarrhea with an estimated annual incidence over 300,000 cases in the United States (1). The primary challenge in treating CDI is recurrence (rCDI) which occurs in as many as 20-35% of patients within 60 days of completing treatment despite a lack of apparent risk factors. Furthermore, the risk of rCDI increases to 33-65% in patients with a history of prior recurrence. Therefore, rCDI is responsible for over 50% of healthcare costs associated with CDI (2-4). This situation has been further compounded by the emergence of more aggressive strains of CD in the last two decades that have increased both the severity of primary infections and the frequency of recurrence (5-7).

Two approaches have been shown to reduce CDI recurrence rates. Bezlotoxumab (commercially, Zinplava™), a monoclonal antibody therapy directed against *C. difficile* toxin B launched in 2017, provided 40% protection against recurrence in immunocompromised patients at high-risk and did not appear to interfere with repopulation of the gut microbiome (2, 8-13). The second is fecal microbiota transplantation (FMT) wherein the antibiotic-depleted gut flora is repopulated with the microbiome harvested from healthy donors. Used primarily for patients who have suffered multiple recurrences, FMT has proven remarkably successful (14-24). However, these strategies are reserved for high-risk primary CDI patients, especially those with multiple recurrences; cost precludes their broader use in most cases of primary CDI since the majority of patients do not recur after the first episode. Therefore, a diagnostic test that can identify patients at-risk for recurrent disease after the first episode would facilitate the adjunctive use of these therapies.

We have recently developed a novel immunoassay, MENSA (Medium Enriched for Newly Synthesized Antibodies), in which specific antibody production by recently activated antibody secreting cells (ASCs) is measured in response to primary CDI (25). During an acute infection, activated B cells in lymph nodes proliferate and differentiate into ASCs that rapidly appear in the circulation for days. In the case of CDI, the majority of ASCs produce CD-specific antibodies before they undergo apoptosis (26). A small fraction of ASCs migrates to secondary lymphoid organs and the bone marrow to become long-lived plasma cells (27). Although the role of antibodies in the serum to CDI has been debatable in some studies (28-31), we show ASCs in MENSA offer immune correlates of protection. In all, *Clostridioides difficile (CD)-*specific antibodies made by ASCs in the MENSA measures active immune responses to identify patients at low-risk for recurrence after primary CDI. Importantly, the timing of the MENSA response is crucial, as the appearance of ASC can precede seroconversion by a few days and provides earlier identification of patients at low risk for recurrence (26).

In this study, we examine the MENSA and serum responses to ten CD antigens in 46 primary CDI patients and show that IgA and IgG responses to three of those antigens provide a simple diagnostic tool for predicting non-recurrence.

## RESULTS

### Human Subjects

After approval from Emory University, Dekalb (now Emory Dekalb) and Grady Institutional Review Boards, patients with primary CDI were enrolled and followed for recurrence. The primary CDI population (n=46) was similar in size to the control population (n=64) and in the proportion of races (*p*=0.52) and sexes (*p*=0.21; Table 1) (25). The control population was younger than the CDI population (*p*<0.0001), but there was no significant difference between younger and older healthy controls in serum and MENSA levels of antibodies specific for *C. difficile* antigens (MENSA *p*=0.71; Serum p=0.07, older subjects slightly lower). More importantly, the recurrent CDI population (n=9) and the non-recurrent population (n=37) were similar in terms of age (over 50 years of age; *p*=0.46), race (∼60% black; ∼40% white; *p*=0.70), and sex (∼60% female; *p*=1.0). The nine recurrent patients experienced recurrence onset 16-51 days post symptom onset (DPSO; median = 28 DPSO, mean = 30 DPSO); or 4-39 days following the end of initial antibiotic therapy.

**Table 1.** Demographics of control and *C. difficle*-infected subjects

|  | Age |  | Race* |  | Sex |  |
| --- | --- | --- | --- | --- | --- | --- |
|  | < 50 | ≥ 50 | Black | White | Female | Male |
| **C<sub>0</sub> Controls** |  |  |  |  |  |  |
| Healthy (N=38) | 31 (82) | 7 (18) | 19 (50) | 10 (26) | 26 (68) | 12 (32) |
| Medical (N=26) | 22 (85) | 4 (15) | 19 (73) | 6 (23) | 21 (81) | 5 (19) |
| Total (N=64) | 53 (83) | 11 (17) | 38 (59) | 16 (25) | 47 (73) | 17 (27) |
| **CDI Patients** |  |  |  |  |  |  |
| Non-Recurrent (N=37) | 14 (38) | 23 (62) | 23 (62) | 12 (32) | 22 (59) | 15 (41) |
| Recurrent (N=9) | 2 (22) | 7 (78) | 5 (56) | 4 (44) | 6 (67) | 3 (33) |
| Total (N=46) | 16 (35) | 30 (65) | 28 (61) | 16 (35) | 28 (61) | 18 (39) |
| C<sub>0</sub> Controls vs CDI Patients | $p < 0.0001$ | | $p = 0.52$ | | $p = 0.21$ | |
| Non-Recurrent vs Recurrent | $p = 0.46$ | | $p = 0.70$ | | $p = 1.0$ | |

\*Race percentages < 100% due to small sub-population of other races not listed.

### Sample collection

Whole blood samples were drawn during acute infection (2-12 DPSO) and recovery/recurrence periods (13-60 DPSO) to determine the timing of MENSA and serum responses. Because CDI is defined as watery diarrhea three or more times per day for 2 days, most patients began treatment on 2 DPSO. Antibiotic treatment typically lasted 10 days, so the clinically relevant interval for prediction of non-recurrence was within 12 DPSO.

### *C. difficile* antigens in MENSA and serum assays

The combined antibody responses (IgA+IgG+IgM) specific for ten *C. difficile* antigens including: TcdB-CROP, TcdBvir-CROP, CDTb, TcdA-CROP, Cwp84, FliC, TcdB-GTD, GDH, CDTa, and TcdA-GTD, were measured as previously described (25). Among the 46 CDI patients who provided samples 2-12 DPSO, the 9 recurrent patients made little or no anti-CD antibody in their MENSA samples whereas most non-recurrent patients generated new ASCs indicated by their positive MENSA responses (Fig. 1). Twenty-six of the 37 (70%) non-recurrent patients demonstrated a positive MENSA response specific for at least one CD antigen (Fig. 2). Among the ten antigens, responses to four were prominent in terms of: 1) magnitude (percentage of positive patients whose measured antibody levels were greater than five times the C_0_); and 2) frequency of response (percentage of patients who were positive): TcdB-CROP, TcdBvir-CROP, CDTb, and TcdB-GTD. Specifically, among the 26 MENSA-positive, non-recurrent patients, 17 were positive for TcdB-CROP, 19 for TcdBvir-CROP, 12 for CDTb and 14 for TcdB-GTD.

**Figure 1.**
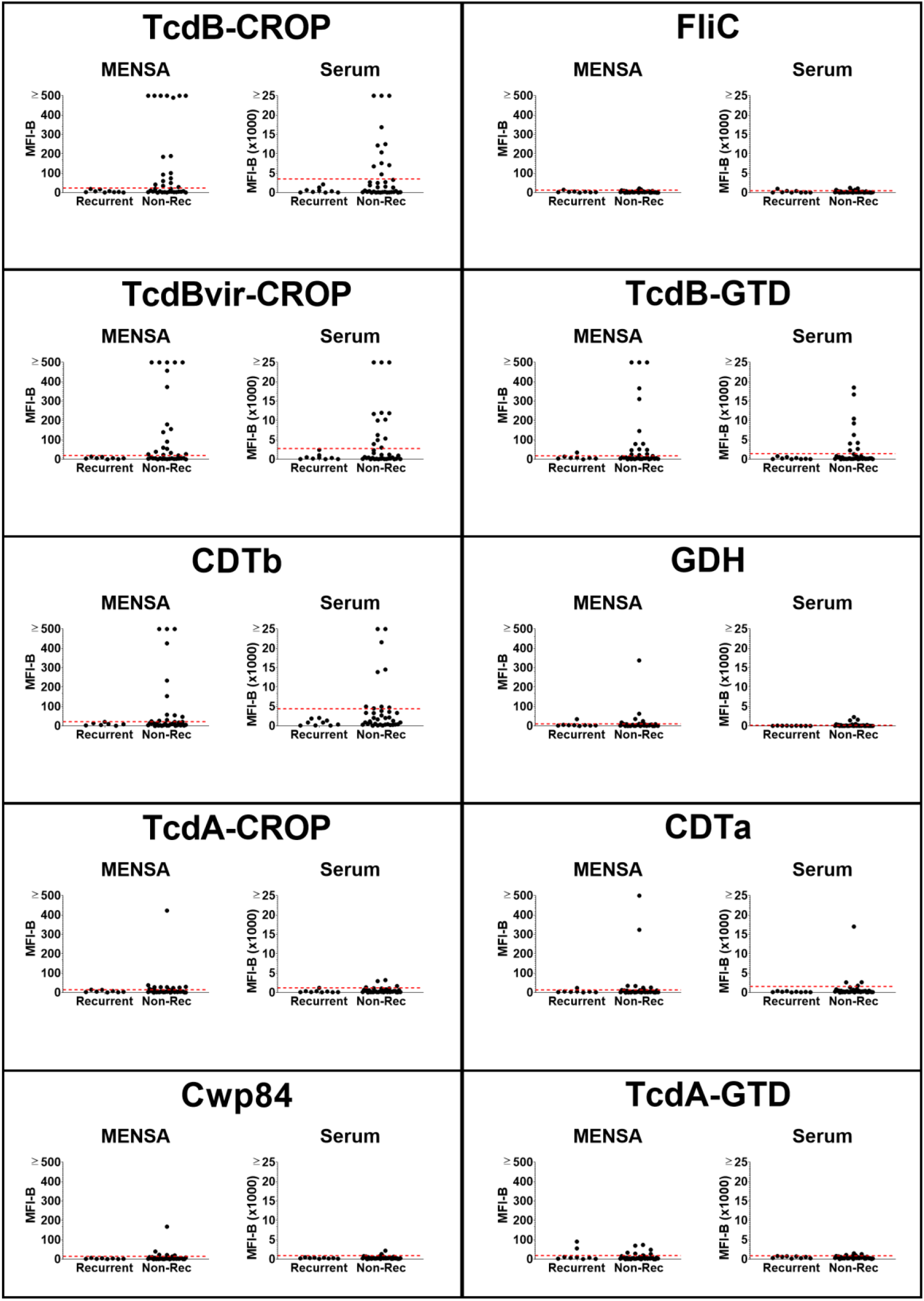
Antibody levels in MENSA and serum samples specific for 10 CD antigens are found at greater frequencies and magnitudes in non-recurrent CDI patients compared to those in patients who will experience recurrence. Levels of antibodies against each of the 10 CD antigens in MENSA and serum samples 2-12 DPSO prepared from 37 non-recurrent patients and 9 recurrent patients are compared. Measured values from each patient (black dots) against each antigen (listed at the top of each of the ten segments in the figure) are presented separately with the MENSA responses on the left and serum responses on the right; measured responses on samples from recurrent patients are on the left in each panel and those from the non-recurrent patients are on the right. C_0_ threshold values, based on control samples prepared from medical workers and healthy adults, are represented by dashed red lines (25). MFI-B values represent the sum of IgA, IgG and IgM antibodies reactive against each of the 10 antigens. Scale for MENSA samples ranges from 0 to 500 MFI with background subtracted (MFI-B; background MFI typically ∼10 for samples run undiluted); scale for serum samples runs from 0 to 25,000 MFI-B on samples diluted 1:1000.

**Figure 2.**
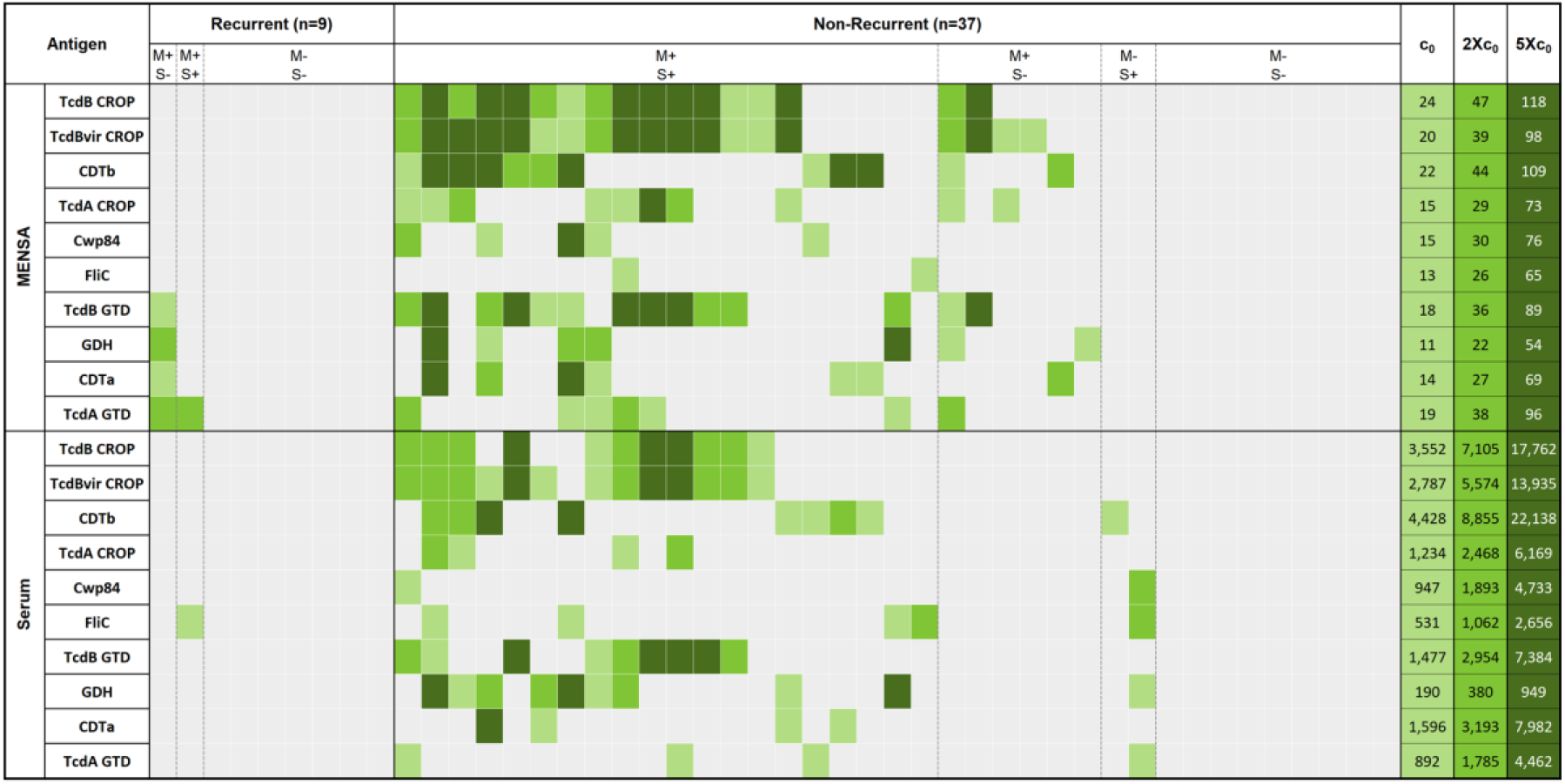
Non-recurrent patients had elevated levels of anti-*C. difficile* antibodies in MENSA and serum. Analysis of samples drawn 2-12 DPSO yielded this heat map showing the responses of each recurrent (n=9) and non-recurrent patient (n=37). Each patient’s responses to the ten CD antigens (listed on the left) are presented in a single vertical column, MENSA levels on the top and serum levels on the bottom. Recurrent and non-recurrent patient groups are further subdivided into groups with antibodies present in: 1) both MENSA and serum samples (M+S+); 2) MENSA samples only (M+S-); 3) serum samples only (M-S+); and 4) neither MENSA nor serum (M-S-). MFI-B values less than C_0_ are presented in grey; the values for C_0_, 2XC_0_, 5XC_0_ are listed at the extreme right of the figure with their respective shade of green. C_0_ values were determined using samples prepared from the population of 64 healthy controls and healthcare workers in Table 1 (25).

Serum antibody responses from most patients were similar to those measured in MENSA samples collected on the same days (Figs. 1, 2). Among the 37 non-recurrent patients, serum antibody levels identified 59% (22/37) of those with ongoing *C. difficile* infections. Eleven were positive for TcdB-CROP, 13 for TcdBvir-CROP, 9 for CDTb, and 9 for TcdB-GTD. The other six antigens elicited weaker MENSA or serum antibody responses from smaller subsets of patients (Fig. 1, 2).

### Antigen selection

Three secreted toxin antigens (TcdB-CROP, TcdBvir-CROP and CDTb) were the most effective for discriminating non-recurrent from recurrent patients in both MENSA and serum (Fig. 3). Two of the recurrent patients had low antibody levels in their MENSA against other *C. difficile* antigens: TcdB-GTD, GDH, CDTa, and/or TcdA-GTD (Figs. 2, 3A); one recurrent patient had a low serum response against FliC (Figs. 2, 3B). With a single exception, non-recurrent patients who secreted antibodies specific for TcdB-GTD also produced antibodies against the 3 toxin-derived antigens (TcdB-CROP, TcdBvir-CROP, and CDTb). In combination, MENSA responses to these three antigens positively identified the majority of MENSA-positive, non-recurrent patients (23/26; 88%). Similarly, antibody responses in serum against the same three antigens identified the majority (19/22; 86%) of those positive for any CD antigen. Therefore, we focused the analyses on the 3 secreted toxin antigens TcdB-CROP, TcdBvir-CROP and CDTb in subsequent experiments.

**Figure 3.**
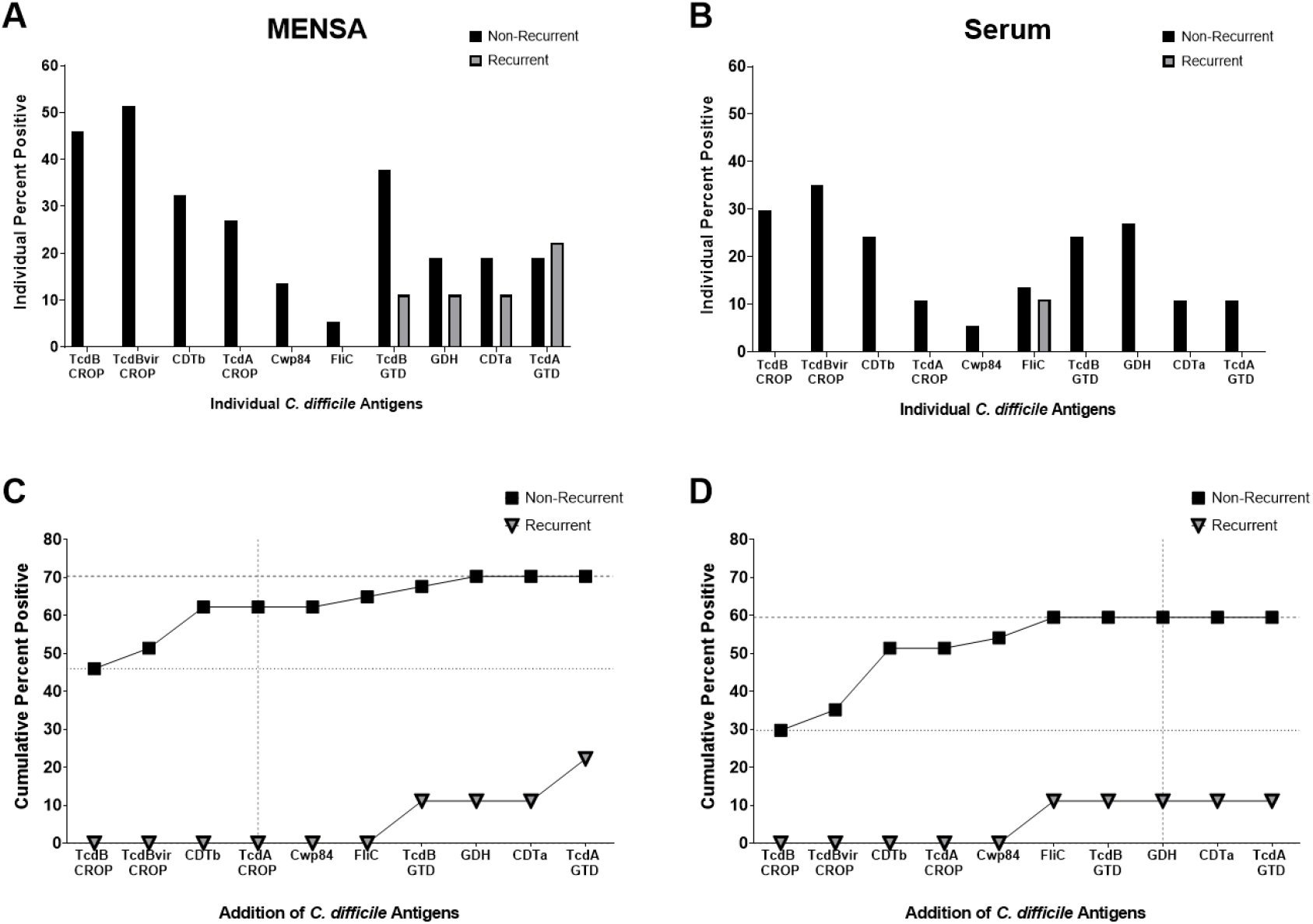
Selection of antigens by maximizing true positive responses and minimizing false positive responses. A) The percentage of non-recurrent (black bars; n=37) or recurrent (gray bars; n=9) patients exhibiting a positive antibody response to each antigen in their MENSA sample. B) The percentage of non-recurrent (black bars; n=37) or recurrent (gray bars; n=9) patients exhibiting a positive antibody response to each antigen in their serum sample. C) Combining MENSA antibody responses to certain antigens increases the prediction of true non-recurrent positives while responses to other antigens increase the number of false positive. Percent of patients positive for at least one antigen proceeding from left to right; non-recurrent patients are presented in black squares; recurrent patients are presented in inverted gray triangles. D) Similar analysis for the measurement of antibody levels in serum.

### MENSA and Risk of recurrence

Among the 23 (50%) patients who were MENSA-negative in 2-12 DPSO, nine (∼39%) suffered recurrence in the following six weeks. Thus, patients with a negative MENSA response against all three *C. difficile* toxin antigens had a 19-fold greater relative risk of recurrence compared to MENSA-positive patients (*p*=0.041; Fisher’s Exact Test *p*=0.001). Comparable analysis of serum samples from the same patient population yielded similar results with lower relative risk of 13.6 (*p*=0.067; Fisher’s Exact Test *p*=0.006). Thus, a positive MENSA response correlated with protection from rCDI.

### Kinetics of MENSA and serum responses in CDI patients

To assess the kinetics of antibody responses in MENSA compared to those in sera, percentages of non-recurrent patients positive for the 3 selected toxin antigens in MENSA and serum samples were measured during four successive time intervals: 2-6, 7-12, 13-40, and 41-60 DPSO (Fig. 4A-C). Among the non-recurrent patients, more were positive in their MENSA than in their serum between 2-6 and 7-12 DPSO. In addition, for two antigens, the MENSA compared to serum remained higher up to 40 DPSO. In contrast, the serum responses were better predictors of non-recurrence after day 40.

**Figure 4.**
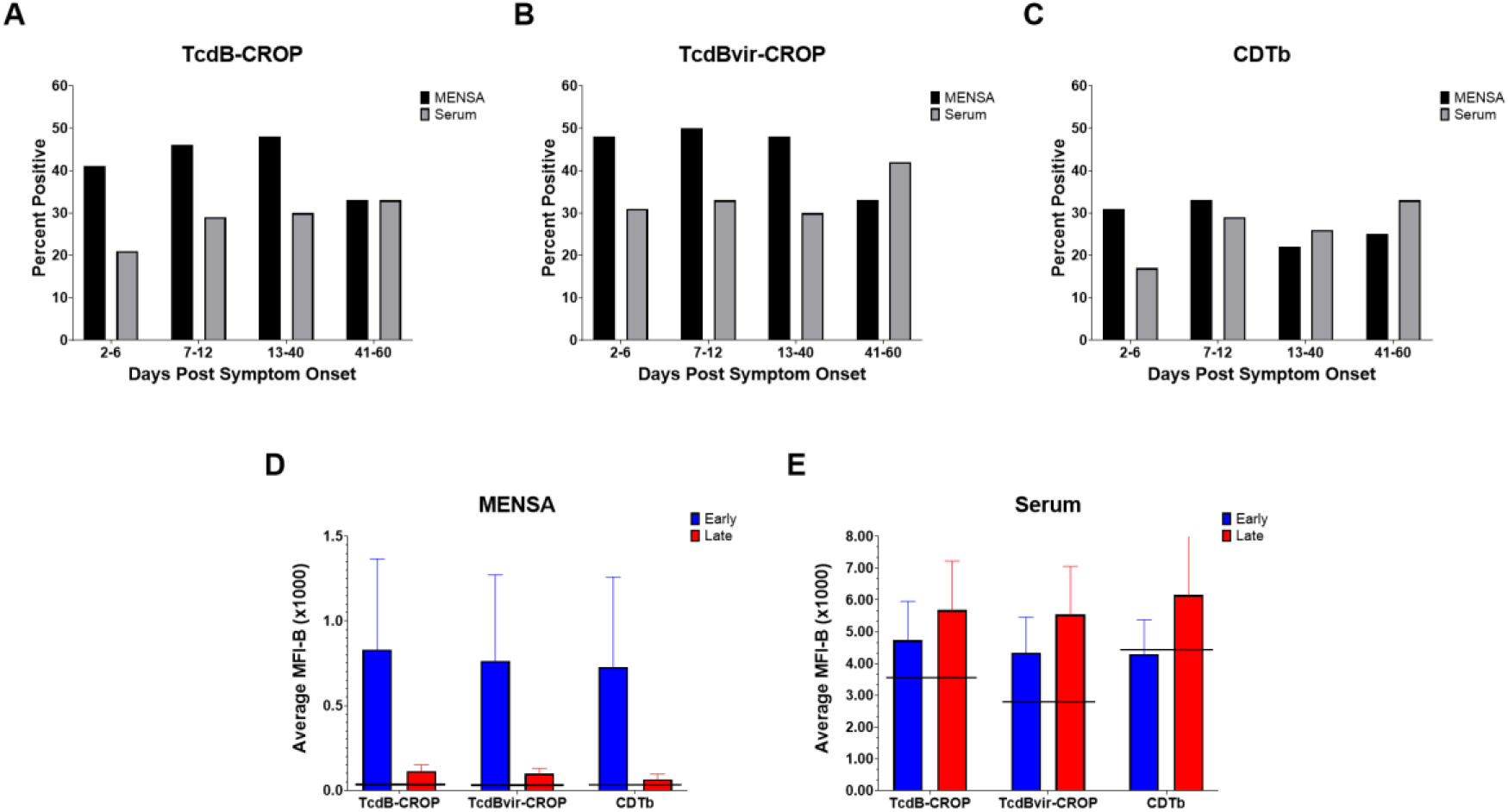
Kinetics of ASC-expressed anti-CD antibodies reveal a peak MENSA response in non-recurrent patients at 2-12 DPSO; serum responses are stronger after 12 DPSO. A) The percentage of patients who had positive anti-TcdB-CROP antibody levels in their MENSA (black) or serum (grey) during different time intervals post-symptom onset: days 2-6 (n=29); days 7-12 (n=24); days 13-40 (n=23); and days 41-60 (n=12). Please note the number of patients in each time interval varies due to timing of sample acquisition. B-C) The same analysis on the same population for antibody levels in MENSA and serum against the other toxin antigens: TcdBvir-CROP (B) and CDTb (C). D) Average levels (MFI-B) of antibodies specific for TcdB-CROP, TcdBvir-CROP and CDTb in MENSA samples from non-recurrent, MENSA-positive patients drawn during days 2-12 (blue) and days 13-60 (red). E) Average levels (MFI-B) of antibodies specific for TcdB-CROP, TcdBvir-CROP and CDTb in serum samples from non-recurrent, MENSA-positive patients drawn during 2-12 DPSO (blue) and 13-60 DPSO (red). Horizontal black lines indicate the C_0_ values for each antigen in MENSA or serum.

The magnitude (MFI-B) of the MENSA and serum responses were compared early (2-12 DPSO) and late (13-60 DPSO) (Fig. 4D-E). *C. difficile*-specific antibody levels declined in the MENSA dramatically at the later time points illustrating the number of circulating ASCs decrease in the blood presumably entering mucosal or bone marrow sites (Fig. 4D). The trend in serum was as expected where antibody levels rose in the first 12 days and remained elevated throughout the 60-day observation period (Fig. 4E).

### High MENSA levels at early time points correlate with later serum antibody changes

A critical element of this model is that the cells responsible for the secreted antibodies (ASCs) emerge into the circulation earlier than the effective rise in serum antibody levels. To examine this hypothesis, the late (13-60 DPSO) increases in serum anti-*C. difficile* toxin levels were plotted against the early (2-12 DPSO) MENSA antibody responses (Fig. 5). Among the 13 patients with positive MENSA and serum titers (NR M+S+; green dots), 9 had an early MENSA response and significant elevation in serum levels later. Two MENSA-positive patients had high serum titers at the early time point and did not demonstrate a rise in serum antibody level (green dots, lower right quadrant). In contrast, the lower left quadrant is populated by patients who had a negative MENSA response and no subsequent increase in serum titers (M-S-). Significantly, this population included the recurrent patients for whom samples were available (Rec M-S-; inverted grey triangles).

**Figure 5.**
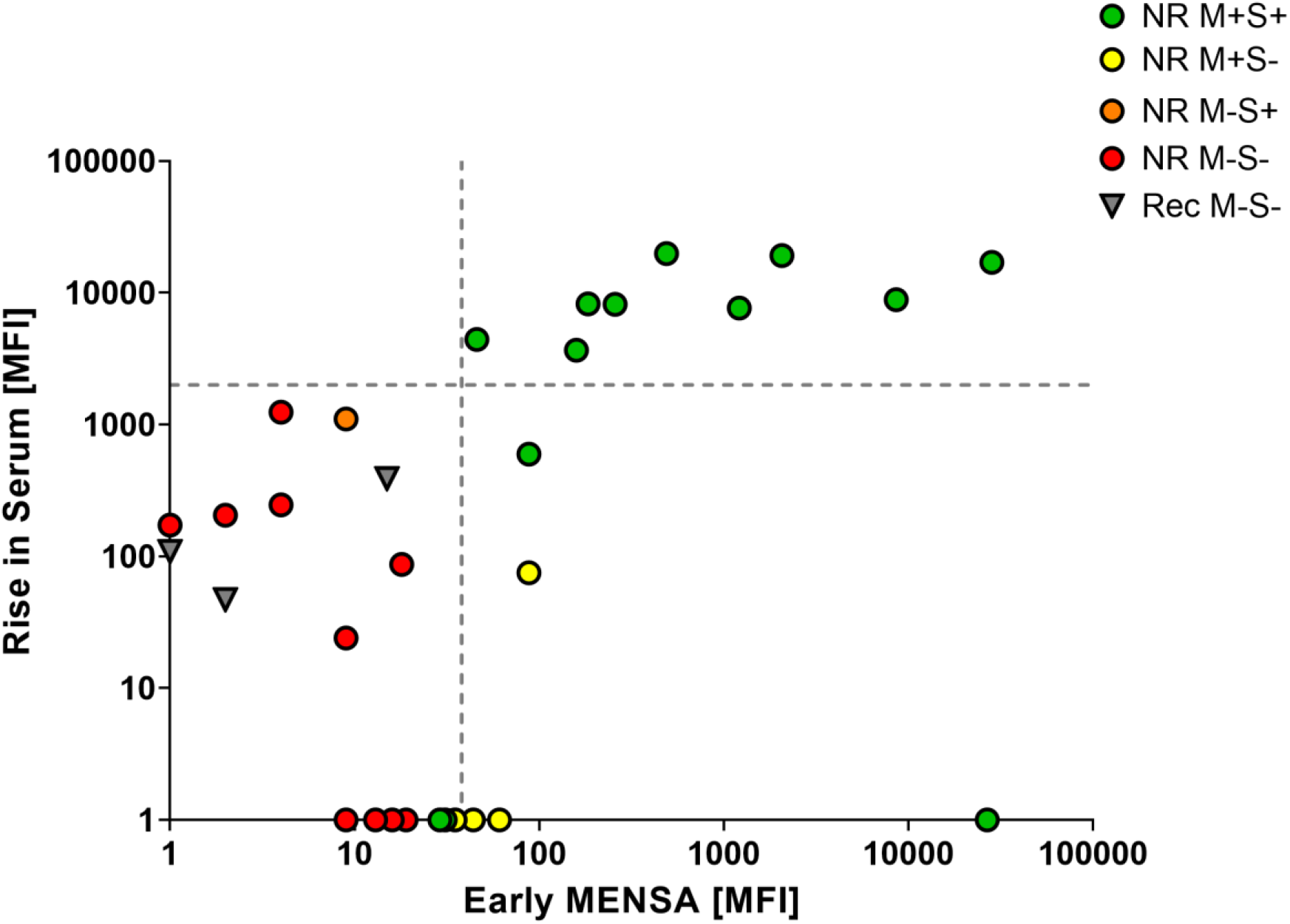
Early MENSA positivity predicts later serum antibody increases. The sum of the MFI-B values for CDTb plus the average of those for TcdB-CROP and TcdBvir-CROP was calculated for each patient at each timepoint. The rise in serum antibodies specific for the three toxin antigens between the first blood draw (2-12 DPSO) and a later blood draw (13-60 DPSO) is presented as a function of the MENSA value at the earlier timepoint. Patients were identified as MENSA and/or serum positive according to Fig. 2. Green circles indicate non-recurrent patients who were positive in their MENSA and serum (NR M+S+); red circles indicate non-recurrent patients who were negative early in their MENSA and never seroconverted (NR M-S-); inverted grey triangles represent recurrent patients (Rec M-S-). The single orange circle represents the only patient in this cohort who was marginally seropositive and MENSA negative (NR M-S+); the yellow circles represent patients who were MENSA positive early but never seroconverted (NR M+S-). The sum of CDTb plus the average of TcdB-CROP and TcdBvir-CROP MFI-B values were calculated for each of the 64 C_0_ control subjects; the vertical dashed line represents the average plus five standard deviations of the C_0_ control population. The horizontal dashed line indicates a 2,000 MFI increase in serum levels between the early (2-12 DPSO) and the late (13-60 DPSO) draws. Due to the log scale of this graph, patients with negative or zero values were placed directly on the x- or y-axis for data inclusion.

### Anti-*C. difficile* antibodies in MENSA are predominantly IgA and IgG

To examine the isotypes of antibodies produced among CDI responses, the levels of Ig against each of the three *C. difficile* antigens were detected with secondary antibodies specific for IgA, IgG or IgM. In serum, IgG predominates (>75% of total Ig). In contrast, IgA and IgG were similarly abundant for the three toxin antigens in MENSA (40-60%), with IgM contributing less than 10% (Fig. 6A-B).

**Figure 6.**
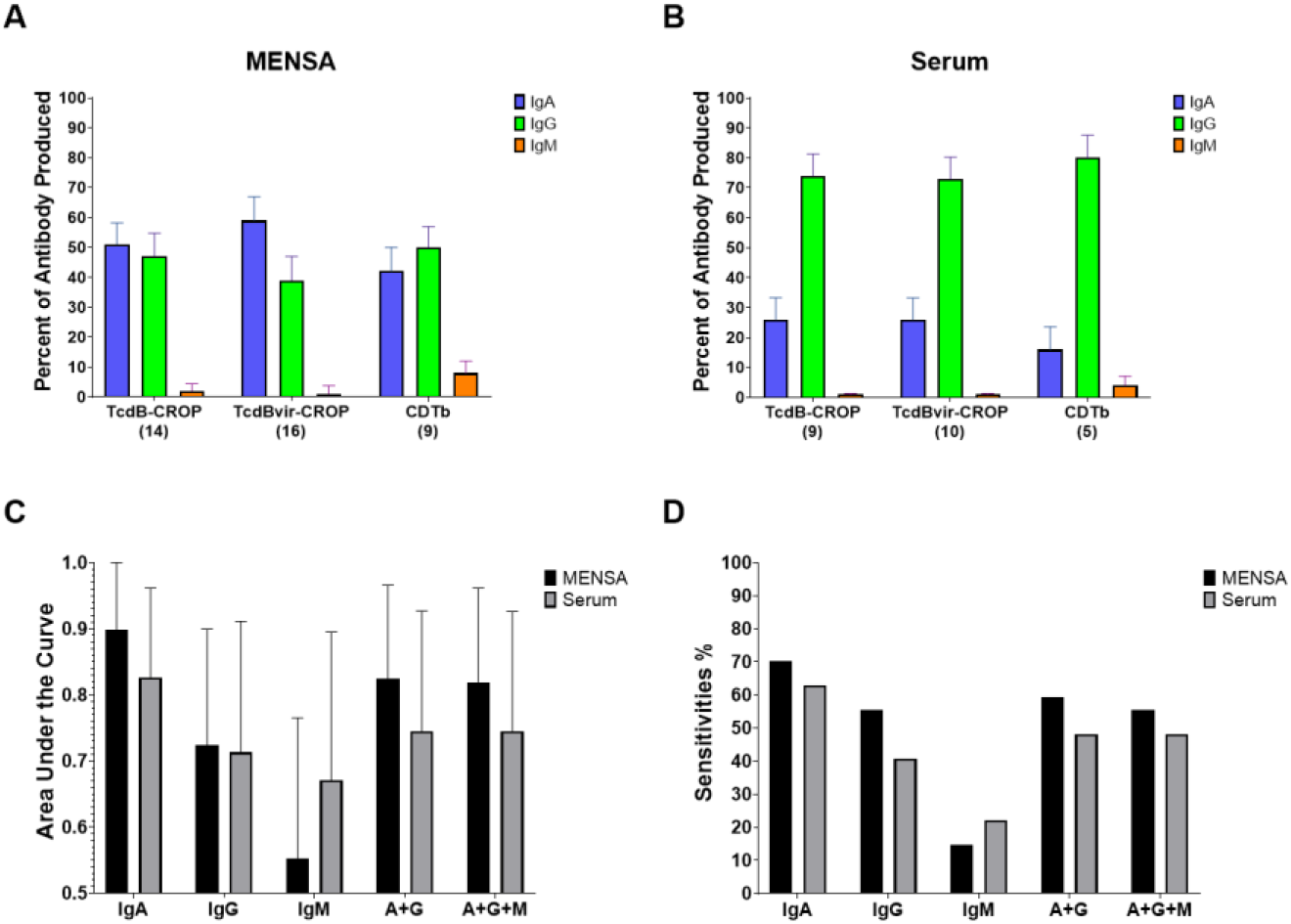
Elevated MENSA and serum IgA, IgG, or IgM levels of anti-TcdB-CROP, anti-TcdBvir-CROP and anti-CDTb. A) Bar graph showing the relative abundance of the different immunoglobulin isotypes in the MENSA response to each antigen. Each antigen is indicated on the x-axis; the number beneath the antigen indicates the number of patients who were positive for that antigen in total Ig and for whom sufficient sample remained for analysis. The y-axis (percent of antibody produced) indicates the average percentage of the total specific antibody response in each positive patient sample that was of the indicated Ig isotype: IgA (blue); IgG (green); IgM (orange). B) The same analysis applied to serum samples. C) Area-under-the-curve (AUC) values determined by ROC curve analysis of CDTb plus the average of TcdB-CROP and TcdBvir-CROP from the 35 (n=27 for non-recurrent; n=8 for recurrent) patients in this study for whom sufficient sample remained for separate isotype analysis. AUC values range from 0.5 (no predictive value) to 1.0 (perfect predictive value); Black bars indicate AUC values for MENSA samples; Grey bars indicate AUC values for serum samples. Error bars indicate 95% confidence interval upper limit. D) Sensitivity values from the same ROC analysis in C.

### IgA responses in MENSA correlate with recurrence-free recovery in the first two months

MENSA and serum samples from non-recurrent (n=27) and recurrent (n=8) patients were re-examined with separate detection for anti-IgA, anti-IgG, and anti-IgM for the 3 *C. difficile* toxins and analyzed using receiver operating characteristic curves (ROC). Bar graphs showing AUC and y-axis intercept values (as a minimal measure of analytic sensitivity) for both MENSA and serum are presented in Fig. 6C-D. In MENSA, the AUC values for the resulting ROC curves varied from 0.55 for IgM to 0.90 for IgA. MENSA yielded higher AUC values and sensitivities than serum for all isotypes and combinations except for IgM. The correlation with patients unlikely to suffer recurrence ranged from 15% sensitivity for IgM to 70% for IgA. IgG alone or IgG+IgA yielded AUC values of 0.72 and 0.82 and y-axis intercepts of 56% and 59%, respectively. Analysis of serum samples demonstrated similar trends: IgA alone yielded the highest AUC value (0.83) and sensitivity (64%), followed by combinations of IgA+IgG and IgA+IgG+IgM (AUC=0.75; 48% sensitivity for each), IgG alone (AUC=0.71; 41% sensitivity), and IgM (AUC=0.67; 22% sensitivity). By using MENSA IgA alone or a combination of IgA+IgG, a substantial sub-population unlikely to experience recurrence was identified.

## DISCUSSION

### Summary

Despite medical advances in CDI treatment, the high incidence of recurrence remains a significant unmet need. To address this problem, we demonstrate the diagnostic utility of measuring anti-*C. difficile* antibodies in serum and MENSA. MENSA is a novel analytic fluid containing antibodies produced in vitro by circulating ASC. Unlike serum antibodies, MENSA antibodies reflect only the new humoral response. We found that circulating ASC are present in the blood 2-12 DPSO, and that MENSA antibodies can serve as early biomarkers for protection against recurrence, prior to seroconversion. From a repertoire of 10 CD antigens, we identified three against which MENSA IgA and/or IgG responses were predictive of non-recurrence. Antibodies specific for these three *C. difficile* antigens in the MENSA identified 62% (23/37) of primary CDI patients who would not suffer recurrence.

### MENSA anti-TcdB-CROP and anti-CDTb are early biomarkers of protection against CDI recurrence

Typically, stool samples are tested for *C. difficile* DNA or antigens and then antibiotics are administered for 10-14 days (1). The mean interval between CDI symptom onset and treatment initiation was <2 days for hospital-acquired infection (32). The optimal window for measuring the CDI MENSA response is 2-12 DPSO when most hospitalized CDI patients undergo treatment. Although seroconversion of anti-CD IgG can occur by 12 DPSO (33), our results show that serum antibody measurement can detect up to 50% of *C. difficile*-infected patients within 12 days. In contrast, measurement of antibody responses in MENSA can identify more non-recurrent patients earlier, possibly prior to hospital discharge (26). Most patients with early positive anti-TcdB-CROP, anti-TcdBvir-CROP and/or anti-CDTb MENSA responses developed increased protective serum levels in the weeks following.

### Antibodies in MENSA are better for identification of non-recurrent patients than antibodies in serum

The improved sensitivity in MENSA samples is primarily due to MENSA-positive patients who remained seronegative (Fig. 2, M+S-), an unexpected sub-population. None of the six patients in this group experienced recurrence, a group too small to assess clinical significance (p=0.27). Exactly what defines this sub-population is not known; however, MENSA responses may arise without seroconversion if the immune response is aborted early or if the circulating IgA-ASC migrate to mucosal sites and do not contribute to the serologic response. Thus, MENSA alone could be an effective biomarker for protection from recurrence.

### The secreted toxins are the best antigens for diagnosis of non-recurrence

For this test, of the *C. difficile* antigens, the secreted toxins, TcdB-CROP, TcdBvir-CROP and CDTb provided the best results. Although *C. difficile* displays an array of cell surface and secreted antigens and extensively described in our previous studies (25), a defensive enzymes (GDH (34)), a flagellar components (FliC (35-37)) and a cell wall protein associated with biofilm building (Cwp84 (38-41)) added little or no value for MENSA or serum-based diagnostics. Antibody levels specific for the TcdA domains, CROP and GTD, were redundant with the larger responses to the TcdB and CDTb toxins. Likewise, the TcdB-GTD and CDTa antigens added little or nothing. Thus, these antigens were eliminated from further consideration.

### IgA is a strong predictor in both MENSA and serum

In serum, the majority of measured antibodies were IgG (73-80%), while in MENSA, IgG and IgA were equally abundant (39-50% and 42-59%, respectively). In both MENSA and serum samples, IgA alone yielded higher AUC values and sensitivities than the other isotypes alone or in combination suggesting limiting the MENSA measurement to IgA alone. As a pathogen of the gastrointestinal tract, *C. difficile* is likely to elicit a strong IgA response. However, IgG had greater magnitude in serum and MENSA responses demonstrating its importance. Additionally, our patient population was biased towards TcdB-CROP-positive strains due to the PCR test used for screening. Given a cohort with more binary toxin-positive patients, IgG may be of greater utility. Therefore, we conclude that the MENSA immunoassays should include both IgA and IgG responses to these three *C. difficile* antigens.

### Limitations of this study

Fundamental limitations of this study were the population size and enrollment of patients from a tertiary referral center with a substantial number of immunocompromised patients due to transplantation and malignancies. Enrollment from community hospitals that reflect the primary CDI burden in United States healthcare settings would improve future studies.

Another concern was the utility of TcdB gene PCR in stool samples for primary diagnosis for this study. Recent reports suggest that screening for CD genes by PCR may overestimate the frequency of infections (42, 43), due to its inability to distinguish true infections from colonization (44, 45). Since colonization does not elicit a MENSA response, some of our MENSA-negative patients may not have had true CDI.

Finally, two of the selected antigens are the secreted toxin TcdB-CROP domains produced by the historical strain VPI 10463 and the hypervirulent epidemic strain, NAP1/B1/027 R20291 (46). Results for MENSA or serum antibodies specific for each antigen are similar but the selection of a single or combination antigen will require additional clinical evaluation.

### MENSA-based diagnostics in primary CDI targets preventative rCDI therapies

Recently, Bezlotoxumab (tradename: Zinplava™) (13) and FMT have been shown to prevent rCDI. The favorable safety profiles of these therapies make a reasonable case for administration after primary CDI in the subset of patients who are at risk for recurrence. However, if given to all patients with primary CDI, only 30% could actually benefit. The utilization of our *C. difficile* MENSA immunoassay at the time of primary CDI could identify patients at the greatest risk of recurrence.

In conclusion, we propose a new strategy for stratifying patients during primary CDI into those at low versus higher risk for recurrence. The model measures the host immune response as a clinical prediction tool for recognizing CDI patients not at risk for recurrence and thereby enabling preventative measures to be directed to patients who would derive maximal benefit. Ultimately, this *C. difficile* MENSA immunoassay could lead to substantial healthcare cost reductions while decreasing recurrence.

## METHODS

### Study Approval

The Emory and Dekalb Institutional Review Boards and the Grady Research Oversight Committee approved all protocols and procedures. Written informed consent was obtained from each patient prior to inclusion in the study.

### Enrollment of CDI patients and controls

CDI patients and control subjects were recruited at Emory University and Dekalb Medical Center (now Emory Decatur Hospital) from 2015-2017. A total of 46 patients with CDI validated by PCR and/or ELISA (Xpert *C. difficile*; C. DIFF QUIK CHEK COMPLETE, Alere) were enrolled. Whole blood samples were collected at one-to-three time points (Draw 1: 1-5 days post-validation (DPV), Draw 2: 6-12 DPV, Draw 3: 21-60 DPV). A patient was considered recurrent if he/she had follow-up diarrhea that was positive by PCR or QUIK CHEK within 60 DPSO. For controls, blood was collected at a single time point from 38 healthy donors, and 26 medical workers from Emory Hospitals. Samples were transported at room temperature (RT) to the MicroB-plex laboratory and processed within 24 hours.

### MENSA generation

PBMC were isolated by centrifugation (800xg; 25 min) using Lymphocyte Separation Media (Corning). Five washes with RPMI-1640 (Corning) were performed to remove serum immunoglobulins (800xg; 5 min), with lysis of residual erythrocytes after the second wash and cell counting after the fourth. Harvested PBMCs were cultured at 10^6^ cells/mL in R10 Media (RPMI-1640, 10% Sigma FBS, 1% Gibco Antibiotic/Antimycotic) on a 12-well sterile tissue culture plate for 24 hours. After incubation, the cell suspension was centrifuged (800xg; 5 min) and the supernatant (MENSA) was separated from the PBMC pellet, aliquoted and stored at -80 °C.

### Serum generation

One clot activator tube containing 2-4mL whole blood was collected and incubated at RT for at least 30 minutes. The clot was discarded, and remaining supernatant was centrifuged (800xg; 10 min), removed from pellet, aliquoted and stored at -80 °C.

### Selection and preparation of recombinant antigens

Selection and design of CD recombinant antigens are presented in detail (25). Briefly, antigens or domains thereof were selected based on known immunogenicity and/or pathogenicity. The antigen repertoire included: 1-4) the combined repetitive oligopeptide (CROP) and glucosyl transferase (GTD) domains of the secretory toxins TcdA and TcdB; 5) the CROP domain of TcdB associated with hypervirulent CD strains (TcdBvir-CROP); 6-7) the A (CDTa) and B (CDTb) subunits of the secreted binary toxin CDT, 8) flagellin (FliC), 9) a major cell wall protein (Cwp84); and the enzyme glutamate dehydrogenase (GDH). Published sequences were examined for homology across CD strains and truncated to remove potentially problematic regions such as signal peptides and cross-reactive domains. His- or GST-tags were added to the N- or C-terminus of each protein to facilitate purification and or expression. Each protein was characterized by size (SDS-PAGE), purity, and immuno-reactivity with available monoclonal antibodies or pooled positive sera from infected patients.

### Multiplex immunoassays for quantification of anti-*C. difficile* antibodies

All samples were tested using a multiplex immunoassay protocol previously described using Luminex MAGPIX® (25). All MENSA samples were tested undiluted; serum samples were diluted 1:1000 in Assay Buffer (PBS, 1% BSA, pH7.5).

### Data analysis

Median Fluorescent Intensity (MFI) of combined detection antibodies (IgA+IgG+IgM) was measured using xPONENT 4.2 software. The background fluorescence of assay buffer or R10 (∼10-30 MFI) was subtracted from each serum or MENSA result, respectively, to obtain MFI minus Background (MFI-B). Positive cut-off values (C_0_) for each antigen were determined as the average plus 5 standard deviations (Avg+5SD) of 64 control subjects in MENSA samples and the Avg+4SD for serum samples. For assay evaluation, patient samples collected within 2-12 DPSO were selected. In patients with two blood samples during this interval, the one with the higher sum of MFI-B values against nine antigens was selected (TcdBvir-CROP was excluded due to its similarity with TcdB-CROP to avoid duplication). Analysis and data representation were undertaken using Microsoft Excel, GraphPad Prism, and JMP statistical analysis packages.

### Statistics

Determination of relative risk of recurrence between those patients positive in their MENSA for anti-TcdB-CROP, anti-TcdBvir-CROP and/or anti-CDTb and those who were antibody-negative was calculated using online statistical software for Relative Risk (MedCalc.org) and Fisher’s Exact test (MedCalc.net).

## Data Availability

All data produced in the present study are available upon reasonable request to the authors

## AUTHOR CONTRIBUTIONS

FL and JD designed and supervised the study. NH, SN, SO, FL, and JD wrote the manuscript with support/input/feedback from all authors. NH, SN, GK, and SO contributed to sample preparation, carried out the experiments, and analyzed the data. CK, AB, and PR acquired the patient samples. YW confirmed CDI diagnosis in patient samples. HW provided statistical guidance. SL oversaw patient recruitment. LC reviewed initial study design and data. MK provided detailed editing and thoughtful review of the manuscript. Each author contributed to the manuscript revision, read and approved the submitted version.

## ACKNOWLEDGEMENTS

This work was supported by the Centers for Disease Control and Prevention through Research Contract #200-2015-88233 titled “Antibody Secreting Cells (ASC) in Human *Clostridium difficile* Infection, Colonization, and Recurrence”. Special thanks are due to the clinical coordinators who assisted in this project. Coordinators at Emory Hospitals were Ms. Hinel Patel, Ms. Aja Bowser and Ms. Maya Lindsay. Our thanks also to the clinical coordinators at Dekalb Hospital and IDS Atlanta, Mr. Travis Stewart and Ms. Julia Norton. Finally, special thanks to our sponsor on this effort, Dr. L. Clifford McDonald at the Centers for Disease Control for his continued advice and support.

